# Impact of Remote Cholecystectomy on Clinical Outcomes Following *Pseudomonas aeruginosa* Bloodstream Infection: A Retrospective Cohort Study

**DOI:** 10.1101/2024.01.26.24301822

**Authors:** Hanna K. Bertucci, Lyndsey R. Heise, Michael P. Angarone, Alan R. Hauser, Marc H. Scheetz, Kelly E. R. Bachta

## Abstract

**Purpose:** Mortality associated with *Pseudomonas aeruginosa* bloodstream infection (PABSI) remains high despite advances in clinical care and therapeutics. In a recent study using mice to model PABSI, the gallbladder (GB) was identified as a reservoir for bacterial expansion. Furthermore, bile exposure has been linked to increased antimicrobial resistance (AMR). Therefore, we asked whether patients with retained gallbladders might suffer from more antimicrobial resistant *P. aeruginosa* (*Pa*) infections, extended culture positivity, and worsened clinical outcomes.

**Methods:** Retrospective cohort study of adults hospitalized over a five-year period with PABSI. PABSI cases were defined as patients with ≥ 1 positive *Pa* bacterial culture from the blood. Patients were categorized as either those retaining a gallbladder (no cholecystectomy) or those without (cholecystectomy). Cholecystectomy was defined as a history of cholecystectomy ≥ 1 year prior to the index episode of PABSI. Inferential statistics were used to identify associations between remote cholecystectomy and antimicrobial resistance profile, length of blood culture positivity, and in-hospital and 90-day mortality.

**Results:** The final study population included 336 patients, 262 (78%) with retained gallbladder and 74 (22%) without. We observed no difference in 90-day or in-hospital mortality between groups. Overall, composite 90-day mortality was 30.1%. Furthermore, no robust differences were observed in the antimicrobial resistance profile of *Pa* isolates between the groups.

**Conclusions:** In our study, neither PABSI AMR pattern nor clinical outcome was affected by remote cholecystectomy. However, we do demonstrate that mortality for patients with PABSI in the modern era remains high despite advances in anti-pseudomonal therapeutics.

## INTRODUCTION

Bloodstream infections (BSIs), as defined by the ability to culture bacteria from the bloodstream of a patient with signs of systemic infection, are a growing threat to public health. It is estimated that over 575,000 cases of BSIs occur annually in North America leading to over 90,000 patient deaths per year [1]. Positive blood cultures in the context of infection have been shown to increase overall mortality risk and BSIs are a leading cause of sepsis, an often-devastating consequence of infection. Hospitalization due to BSI lasts up to 20 days longer than other admissions, leading to significant economic consequences for the patient and increased resource burden on healthcare systems [2]. In patients whose BSI progresses to clinical sepsis, the average length of hospitalization increases by 75% making sepsis the most expensive condition to treat in the hospital [3, 4]. Sepsis is often fatal and those patients who do survive are more likely to have permanent organ damage and physical and mental impairments [4]. Among all bacterial organisms that cause BSIs, Gram-negative organisms are increasing at an alarming rate.

One such organism is *Pseudomonas aeruginosa* (*Pa*). *Pa* is an opportunistic, environmental Gram-negative bacterium, that commonly causes healthcare-associated infections including BSI. *Pa* is the 4^th^ leading cause of BSIs, comprising approximately 4-9% of all episodes [5, 6]. Compared to all other bacterial species, BSI secondary to *Pa* carries a disproportionately high risk of mortality [7, 8] with reported crude mortality rates ranging widely from 20 to 70% [7–11]. A 13-year prospective study that evaluated 2600 patients with BSI and adjusted for antimicrobial resistance (AMR) patterns and comorbidities, found that the mortality for *Pa* BSI (PABSI) was higher than that of all other bacterial pathogens including *Staphylococcus aureus* [7]. PABSI is typically acquired in situ via central venous catheters, or secondarily following dissemination from an invasive infection such as pneumonia or urinary tract infection [11, 12]. Risk factors for PABSI include prolonged hospitalization, neutropenia, liver and biliary disease, immunodeficiency, the presence of an indwelling catheter and prior gastrointestinal (GI) colonization with *Pa* [10, 13–16]. Factors such as delayed effective antibiotic treatment, neutropenia, underlying pneumonia, and more severe disease at time of presentation have been associated with poor PABSI outcomes [9, 17]. Additionally, rising AMR rates worsen the prognosis for PABSI. Multidrug-resistance (MDR) rates for *Pa* are among the highest of all BSI-causing pathogens, occurring in more than a quarter of all isolates [5]. Given the rise in drug resistance, *Pa* is one of the top two pathogens frequently initially treated with inappropriate antibiotic therapy [18], and inadequate or delayed onset of antibiotic therapy has been shown to worsen prognosis [17, 18]. Despite the development of newer antimicrobial combinations to combat rising AMR (e.g. ceftazidime-avibactam and ceftolozane-tazobactam), limited headway has been made in reducing overall PABSI mortality likely related to an incomplete understanding of pathophysiology.

In a recent report, Bachta, et al. used a mouse infection model to better understand PABSI intra-host infection dynamics [19]. Notably, following PABSI, *Pa* was found to traffic to the gallbladder (GB) where the population expanded dramatically and facilitated high levels of excretion of *Pa* into the gastrointestinal (GI) tract. Remarkably, post infection, the gallbladder contained the highest bacterial burden of any organ. Removal of the gallbladder led to dramatic reductions in the fecal excretion of *Pa*, thereby suggesting that the gallbladder may be a previously underappreciated niche for *Pa* expansion. *Pa* was also shown to replicate robustly in bile *in vitro*. In human studies evaluating the causative agents of cholecystitis and cholangitis, *Pa* is among the top five most frequently isolated pathogens [20, 21], and its presence in the gallbladder has been associated with the development of secondary sclerosing cholangitis [22].

The gallbladder acts as an important reservoir for bile, a complex mixture of bile acids, phospholipids, cholesterol, bilirubin, inorganic salts and trace metals [23]. Bile is one of the most inherently bactericidal compounds in the human body, serving to decrease bacterial burden in the GI tract. Several studies have demonstrated that the gallbladder is an important replicative niche for certain bacterial species. It has been shown to facilitate persistent colonization and enhance transmission of *Salmonella typhi*, *Listeria monocytogenes* and *Campylobacter jejuni* [24–27]. Gallbladder epithelial cells serve as the niche for *in vivo* replication of *Salmonella enterica* serovar Typhi [28] and *L. monocytogenes* replicates within the lumen of the murine gallbladder [29]. As in PABSI, mouse models of BSI demonstrated that *L. monocytogenes* also traffics to the liver and gallbladder and is subsequently excreted in the GI tract [26, 30, 31]. To promote survival and GI colonization, bacteria have developed mechanisms that subvert the toxic effects of bile through adaptation and tolerance [24], and many utilize bile as a signal to enhance virulence gene regulation for efficient infection [24]. *L. monocytogenes* expresses a bile salt hydrolase that, when deleted, decreases virulence [32]. Exposure to bile promotes germination of *Clostridioides difficile*, an important cause of infectious colitis in patients. Other mechanisms that promote bile tolerance include induction of biofilm formation and intracellular invasion of gallbladder epithelial cells, a phenomenon thought to promote chronic carriage and GI shedding of *Salmonella* [33]. Critically, bile exposure has been linked to promotion of increased antimicrobial resistance. There is mounting evidence in *E. coli*, *Enterobacter cloacae*, *Klebsiella pneumoniae*, *Acinetobacter baumannii*, and *Pa* that bile exposure stimulates efflux pump expression enhancing the ability of bacteria to survive in harsh environments [24, 34]. For example, bile exposure in *E. coli* increases expression of multidrug-efflux pumps including AcrAB and EmrAB that promote bile tolerance and increase antibiotic efflux [35, 36]. In *S. typhimurium*, an AcrB-like protein that encodes for an efflux pump contributes to resistance to both bile salts and detergents [37, 38]. Bile exposure in *Enterococcus faecalis* induces genes encoding for EmrB/QacA family drug resistance transporters and increases expression of phospholipid synthetase which is linked to daptomycin resistance [39]. Thus, increased exposure to bile may be contributing to rising antimicrobial resistance in pathogenic bacteria. However, how these responses impact overall survival is poorly understood.

There are limited studies investigating the responses of *Pa*, a non-enteric pathogen, to bile. *Pa* replicates robustly in bile, and bile exposure has been shown to stimulate the production of *Pa* virulence factors including genes important in biofilm formation, cell envelope biogenesis, quorum sensing, Type VI secretion, and antimicrobial tolerance [19, 34, 40–42]. In the lungs of patients with cystic fibrosis and structural lung disease, bile aspiration is associated with the early acquisition of *Pa* [43, 44]. Once established in the lungs of such patients, *Pa* is exceedingly difficult to eradicate as ongoing bile exposure secondary to gastroesophageal reflux potentiates adaptations that allow for persistent colonization [41, 45]. Given that *Pa* replicates in bile, traffics to the gallbladder following BSI, and is commonly cultured from patients with gallbladder infections, we questioned if the presence of a gallbladder played a role in PABSI outcome. We hypothesized that the gallbladder might facilitate greater *Pa* exposure to bile and promote the development of antimicrobial resistance. Additionally, we hypothesized that prior cholecystectomy (gallbladder removal) through elimination of an *in vivo* replication niche, may improve outcomes following PABSI. The purpose of this retrospective analysis was to evaluate the impact of cholecystectomy on AMR development and infection outcomes in patients with PABSI.

## MATERIALS AND METHODS

### Study setting, design, and protocol

In this retrospective cohort study, we examined patients with *Pa* bloodstream infections hospitalized at Northwestern Memorial Hospital (NMH) and Prentice Women’s Hospital from July 1, 2014 to July 1, 2019. NMH is an 894-bed academic medical center with a full range of clinical services including a robust immunocompromised patient population. As *Pa* is rarely a contaminant in the bloodstream, patients were considered to have PABSI if ≥1 blood culture was positive for *Pa*. The first positive isolate per patient was included in this analysis. Exclusion criteria included: patients aged less than 18 years, those who had undergone a cholecystectomy within the last 1 year, patients with HIV/AIDS and pregnant women. Patients with polymicrobial bacteremia were included in the study. This project was reviewed and approved by the Northwestern University Institutional Review Board (project no. STU00210641). Inpatient electronic medical records (NMH Enterprise Data Warehouse), pharmacy and microbiology databases were reviewed. Patient demographics included gender, age, and number of days from admission to the first positive blood culture. The presence of the following comorbid conditions was documented: neutropenia, localized solid tumor, metastatic solid tumor, leukemia/lymphoma, myocardial infarction, congestive heart failure, peripheral vascular disease, COPD, chronic kidney disease, diabetes mellitus, CVA/TIA, dementia, hemiplegia, peptic ulcer disease, liver disease, and connective tissue disease. Charlson comorbidity index (CCI) was calculated at the time of bacteremia to estimate illness severity [46]. Charts were reviewed for evidence of cholecystectomy (gallbladder removal) through identification via ICD-9 or ICD-10 code or through absence of a gallbladder on abdominal computed tomography (CT) scan. For patients with a history of cholecystectomy, the approximate procedure date and reason were recorded, if available. Recorded characteristics of BSIs included the suspected source of infection, whether the infection was polymicrobial, and whether the infection was hospital-acquired (nosocomial, onset ≥ 72 hours after admission). The following aspects of antimicrobial therapy were recorded: time to active therapy, length of blood culture positivity, and culture susceptibility to antibiotics spanning seven classes as detailed below. Length of culture positivity was determined by manual chart review. Outcomes included length of blood culture positivity, 90-day and in-hospital mortality if < 90 days.

### Microbiology

Blood cultures from hospitalized patients were processed by the NMH Microbiology Laboratory using the BacT/ALERT® blood culture system (bioMérieux, Marcy l’Etoile, France), with each set consisting of aerobic and anaerobic cultures. Genus and species identification of bacteria was performed via mass spectrometry identification using MALDI-TOF (bioMérieux) or manual biochemical assays when necessary. Susceptibility testing was performed by either the Vitek II system to determine minimal inhibitory concentration (MIC) or Kirby Bauer disk diffusion on Mueller-Hinton agar plates. Antimicrobial agents tested included ciprofloxacin, cefepime, ceftazidime, piperacillin/tazobactam, ceftazidime/avibactam, ceftolozane/tazobactam, meropenem, imipenem, aztreonam, amikacin, gentamicin, tobramycin, colistin, polymyxin B, and tigecycline. MICs and zones of inhibition were categorically interpreted according to Clinical and Laboratory Standards Institute (CLSI, 30^th^ edition) guidelines [47].

### Definitions

Neutropenia was defined as an absolute neutrophil count < 500/mm^3^ [48]. Nosocomial bacteremia was defined as bacteremia acquired ≥ 72 hours after hospitalization. Polymicrobial bloodstream infection was defined as concurrent isolation of *Pa* plus any additional bacterium from the bloodstream at time of the diagnostic blood culture. Active antimicrobial therapy was defined as therapy with at least one agent to which the causative *Pa* strain was susceptible. Specific doses were not reviewed, but all antibiotics were dosed prospectively by clinical pharmacists to optimize efficacy while minimizing safety concerns. Recurrent PABSI was defined as re-isolation of *Pa* from a patient’s bloodstream ≥ 2 weeks from the incident episode provided interval clearance (documented negative blood culture) of the bloodstream between episodes. Infections were categorized as multidrug resistant (MDR) if they were non-susceptible (either intermediate or resistant) to ≥ 1 agent from ≥ 3 classes of antibiotics, extensively drug-resistant (XDR) if they were susceptible to ≤ 2 classes of antibiotics, and pan-drug resistant (PDR) if non-susceptible to all antibiotics tested following standardized criteria [49]. Culture positivity was analyzed in two ways. First, we recorded the total number of daily positive blood cultures documented regardless of follow up clearance (n=336, all patients). For patients with no blood culture follow-up within 48 hours of index culture, the length of culture positivity was recorded as 1 day. Second, we analyzed only those patients for whom there was documented follow-up of blood cultures within 48 hours of the index positive culture (n = 220). For patients with recurrent PABSI episodes, the longest consecutive duration of documented positive cultures was included.

### Statistical analysis

Statistical analyses were performed using Stata v.17.0 (Statacorp, College Station, Texas). Tables were created with program table1_mc. Results were expressed as mean ± standard deviation (SD). Pearson’s chi-squared test was employed for categorical variables and ANOVA for continuous variables (with appropriate degrees of freedom between groups) to calculate *p-* values. Tests were two-tailed, and a *p*-value of ≤ 0.05 was used for statistical significance. Length of culture positivity data lacked a parametric distribution, and thus *p*-values were calculated using the two-sample Wilcon rank-sum (Mann-Whitney) test. Time to event analyses were completed with log rank tests for assessments of equality. Kaplan-Meier curves were generated using GraphPad Prism v9.0.0 (GraphPad Software, San Diego, California, USA).

## RESULTS

### Patient characteristics

During the study period, 352 episodes of *Pa* bloodstream infection were recorded in 336 unique patients (Table 1) of which twelve patients (3.6%) had recurrent PABSI. The mean age of patients in the study was 66.9 years and 64.3% were male. Nosocomial infections represented 20.8% of all infections and 17.3% of the bacteremia episodes were polymicrobial. Of the individual patients evaluated, 262 (78.0%) had a gallbladder at the time of infection (no cholecystectomy) and 74 (22.0%) had had prior gallbladder removal (cholecystectomy), yielding a ratio of approximately 3:1 of patients with and without a gallbladder at the time of their PABSI. Unsurprisingly given the predilection of females for gallbladder disease, fewer male patients had prior cholecystectomy (43.2% vs. 70.2%, respectively; *P* < 0.001). The majority of patients (64.6%) had a Charlson comorbidity Index (CCI) ≥ 5 at the time of their PABSI diagnosis, and the average CCI was 6.4 in patients without a gallbladder and 5.6 in the cohort with a gallbladder (*P* = 0.026). Generally, the presence of most medical comorbidities was not statistically different between cohorts. However, among patients without a gallbladder, a larger percentage had peripheral vascular disease (20.3% vs. 9.2%, respectively; *P* = 0.008) and liver disease (16.2% vs. 5.0%, respectively; *P* = 0.001). A smaller percentage of patients without a gallbladder were neutropenic (8.1% vs. 22.1%, respectively; *P* = 0.007) or had leukemia or lymphoma (13.5% vs. 25.6%, respectively; *P* = 0.029). The mean time to active therapy was statistically different between the two groups (1.4 days for patients without a gallbladder vs. 0.6 days for patients with a gallbladder; *P* = 0.003).

**Table 1:** Demographic and clinical characteristics of the study population.

| Baseline characteristics | All<br>*n* = 336 | Cholecystectomy<br>*n* = 74 | No<br>cholecystectomy<br>*n* = 262 | P-value |
| --- | --- | --- | --- | --- |
|  | *n* (%) | *n* (%) | *n* (%) |  |
| Male | 216 (64.3) | 32 (43.2) | 184 (70.2) | **<0.001** |
| Age, y, mean (SD) | 66.9 (16.3) | 68.3 (18.2) | 66.5 (15.7) | 0.39 |
| Nosocomial infection<sup>a</sup> | 70 (20.8) | 18 (24.3) | 52 (19.8) | 0.40 |
| Polymicrobial infection | 58 (17.3) | 17 (23.0) | 41 (15.6) | 0.14 |
| Charlson comorbidity index<sup>b</sup>, mean (SD) | 5.7 (2.8) | 6.4 (3.0) | 5.6 (2.7) | **0.026** |
| 1-2 points | 32 (9.5) | 5 (6.8) | 27 (10.3) |  |
| 3-4 points | 79 (23.5) | 12 (16.2) | 67 (25.6) |  |
| ≥5 points | 217 (64.6) | 54 (73.0) | 163 (62.2) |  |
| Days to active therapy, mean (SD) | 0.73 (2.1) | 1.40 (3.5) | 0.55 (1.4) | **0.003** |
| Medical comorbidity |  |  |  |  |
| Neutropenic (ANC <500/mm<sup>3</sup>) | 64 (19.0) | 6 (8.1) | 58 (22.1) | **0.007** |
| Localized solid tumor | 50 (14.9) | 10 (13.5) | 40 (15.3) | 0.71 |
| Metastatic solid tumor | 48 (14.3) | 13 (17.6) | 35 (13.4) | 0.36 |
| Leukemia/lymphoma | 77 (22.9) | 10 (13.5) | 67 (25.6) | **0.029** |
| Myocardial infarction | 58 (17.3) | 13 (17.6) | 45 (17.2) | 0.94 |
| Congestive heart failure | 87 (25.9) | 23 (31.1) | 64 (24.4) | 0.25 |
| Peripheral vascular disease | 39 (11.6) | 15 (20.3) | 24 (9.2) | **0.008** |
| COPD | 47 (14.0) | 14 (18.9) | 33 (12.6) | 0.17 |
| Chronic kidney disease | 44 (13.1) | 7 (9.5) | 37 (14.1) | 0.29 |
| Diabetes mellitus | 94 (28.0) | 24 (32.4) | 70 (26.7) | 0.33 |
| CVA/TIA | 52 (15.5) | 13 (17.6) | 39 (14.9) | 0.57 |
| Dementia | 19 (5.7) | 5 (6.8) | 14 (5.3) | 0.64 |
| Hemiplegia | 13 (3.9) | 2 (2.7) | 11 (4.2) | 0.56 |
| Peptic ulcer disease | 10 (3.0) | 1 (1.4) | 9 (3.4) | 0.35 |
| Liver disease | 25 (7.4) | 12 (16.2) | 13 (5.0) | **0.001** |
| Connective tissue disease | 16 (4.8) | 4 (5.4) | 12 (4.6) | 0.77 |

<sup>a</sup>Nosocomial infection was defined as culture positivity after ≥72 hours of hospital stay.
<sup>b</sup>The Charlson comorbidity index (CCI) was computed on the day of the first positive blood culture.

### Antimicrobial resistance profiles of PABSI isolates

To discern if prior cholecystectomy was associated with the AMR patterns of our PABSI isolates, we examined the resistance profiles of each isolate. Of the 336 unique episodes of PABSI, we had antimicrobial resistance data for all but 13 (3.9%) which were excluded from this analysis (Table 2). As mentioned, 12 patients (3.6%) had recurrent PABSI during this study period. For patients with recurrence, antibiotic susceptibility data was assessed for each patient’s first positive blood culture yielding a total of 323 unique isolates. Overall, for those antibiotics for which we had complete data (ciprofloxacin, cefepime, ceftazidime, piperacillin-tazobactam, gentamicin and tobramycin), we saw the highest rates of non-susceptibility to ciprofloxacin (48/323, 14.9%) followed closely by piperacillin-tazobactam (39/323, 12.1%). The percentage of non-susceptible isolates for each of the above antibiotics was generally similar among patients with and without a gallbladder. A greater proportion of isolates in patients without a gallbladder were non-susceptible to piperacillin/tazobactam (17.1% vs 10.7%, *P* = 0.14), gentamicin (12.9% vs. 7.9%, *P* = 0.20), and colistin (41.7% vs. 30.6%, *P* = 0.48), though none of these results achieved statistical significance. Of the 323 isolates, we had meropenem susceptibility data for 282, of which 8.2% were non-susceptible. Very few isolates underwent testing for ceftazidime/avibactam or ceftolozane/tazobactam as the study period was prior to extensive clinical use of these antimicrobial agents. A relatively large percentage of PABSI episodes were characterized as MDR (12.1%) or XDR (4.3%), but there was no statistical difference in MDR or XDR phenotype observed between patients without or with a gallbladder (MDR 12.9% vs. 11.9%, *P* = 0.82; XDR 2.9% vs. 4.8%, *P* = 0.49, respectively). No isolates were pan-drug resistant. Overall, across all seven classes of antibiotics, we did not observe statistically significant differences in AMR patterns that were correlated with the presence of a gallbladder at the time of PABSI diagnosis.

**Table 2:**
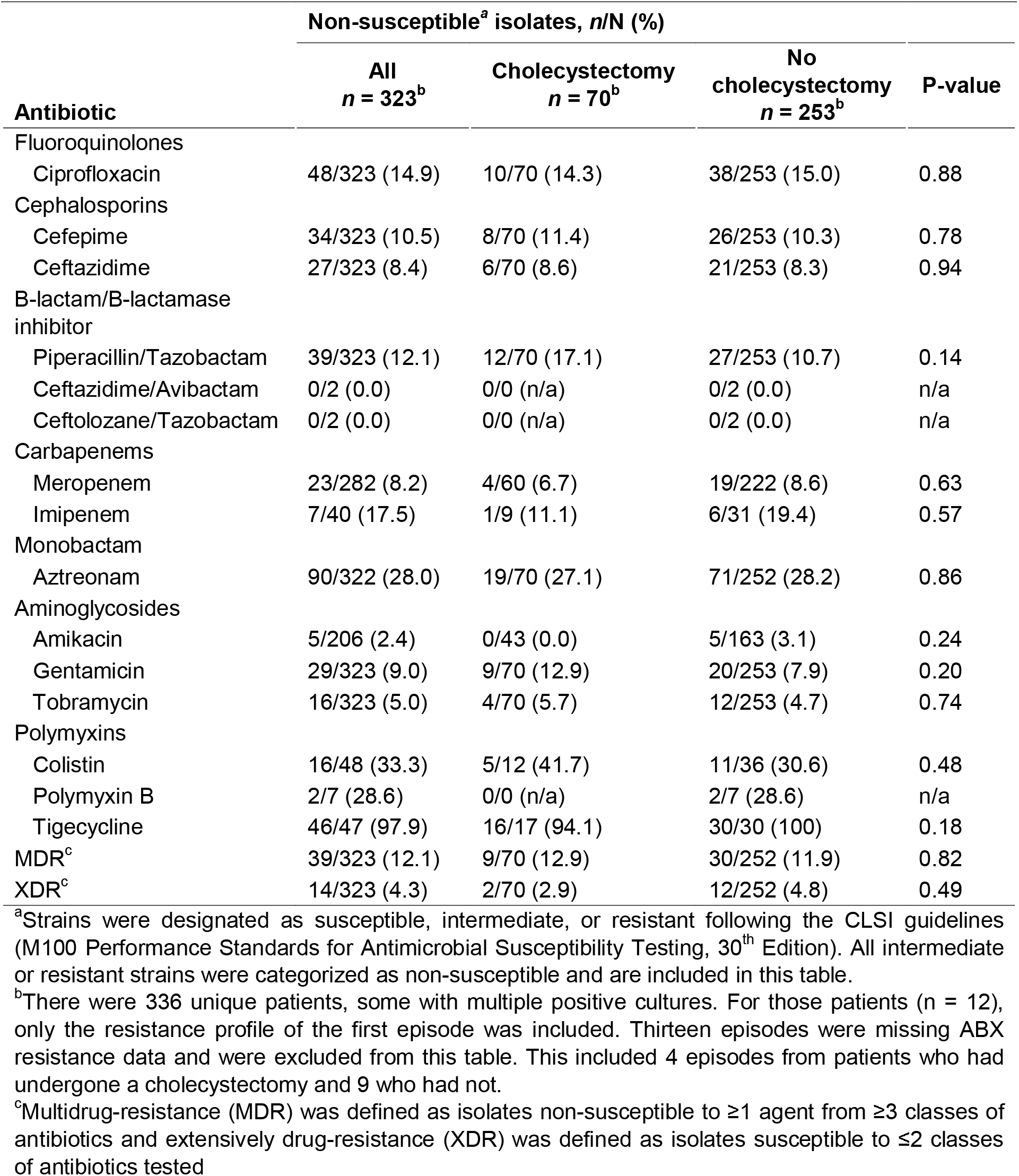
Antibiotic susceptibility patterns in patients with and without prior cholecystectomy

### The impact of prior cholecystectomy on outcome of PABSI

We examined several outcomes measures in our patient population. There was a statistically significant difference in the time to active therapy in the two study populations (1.4 days for the cohort without a gallbladder vs. 0.6 for the cohort with a gallbladder, *P* = 0.003) (Table 1).

No significant difference in duration of bacteremia was observed between the two populations when analyzing all included patients (n = 336). Average length of culture positivity for the entire cohort was 1.2 days, with averages of 1.5 days for the group lacking a gallbladder and 1.2 days for the group who retained a gallbladder (*P* = 0.19) (Table 3). We also analyzed a subset of patients for whom we had documented follow up blood cultures within 48 hours of their index positive culture (n = 220). This subset excluded patients who died prior to repeat blood cultures being drawn. Notably, the majority of patients in both populations (cholecystectomy n = 50, no cholecystectomy n = 170) cleared their cultures within 48 hours. Again, no significant difference in culture positivity was observed between cohorts (1.38 ± 1.2 days vs 1.17 ± 0.6 days, respectively; *P* = 0.44). Ninety-day mortality among both patient cohorts was similar (32.4% and 29.4%; *P* = 0.61), with a considerable number of those patients expiring in the hospital in both cohorts (23% vs. 20.2%; *P* = 0.61) (Table 3). Overall, no statistically significant difference was seen between cohorts for 90-day mortality or in-hospital mortality as seen in Kaplan-Maier curves displaying mortality data (*P* > 0.5) (Fig. 1).

**Fig. 1.**
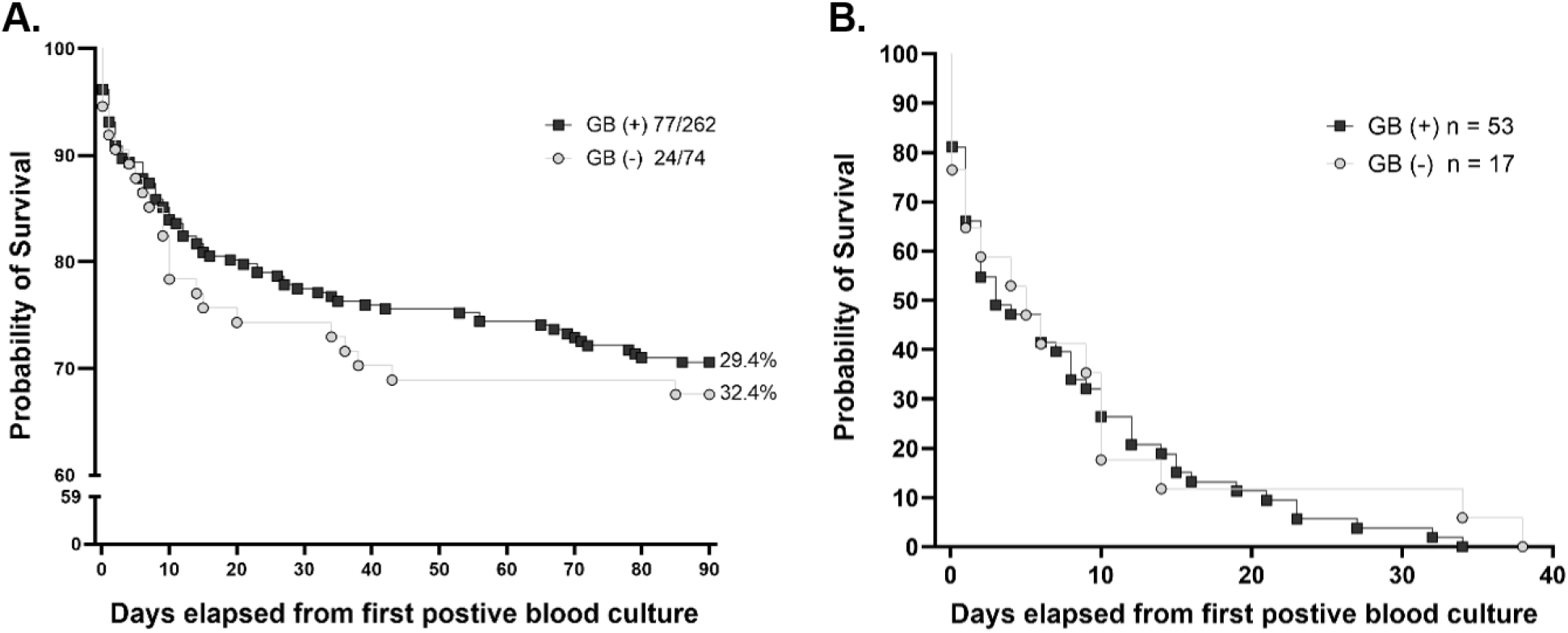
Mortality in Patients with *Pa* Bacteremia. Patients are stratified by history of gallbladder removal (cholecystectomy, gray) or retained gallbladder (black). Results are displayed as a function of 90-day mortality (**A**) and in-hospital mortality (**B**). P-value are >0.5 for both graphs. Day 0 is the day of first positive blood culture

**Table 3:**
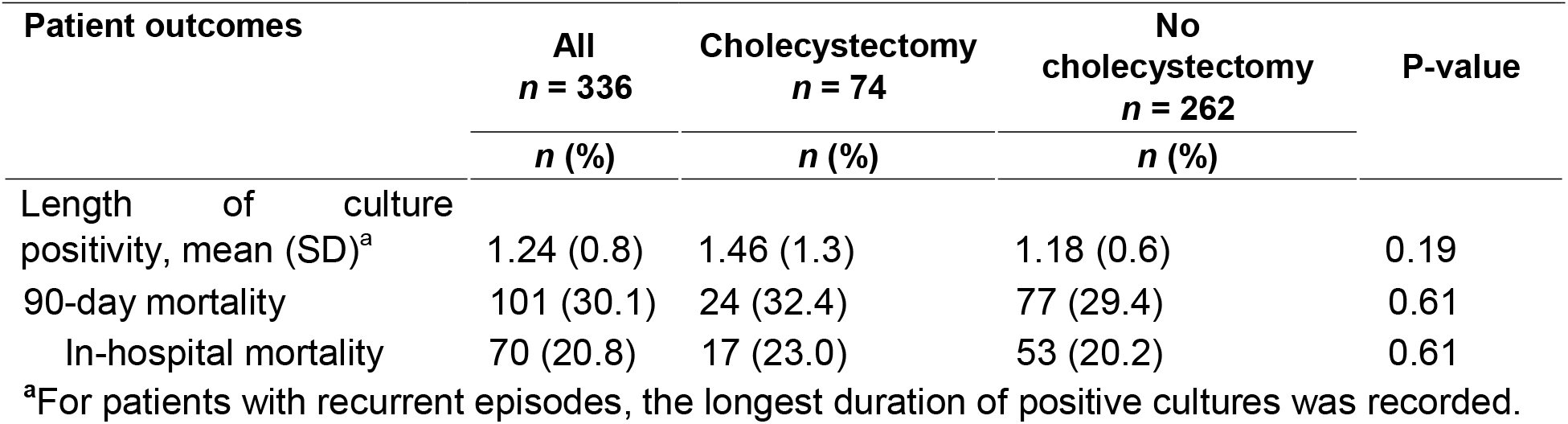
Outcomes in patients with and without prior cholecystectomy

## DISCUSSION

In this single-institution retrospective cohort study, we assessed the impact of prior gallbladder removal on clinical outcomes and *Pa* antimicrobial resistance profile following PABSI. No previous studies have investigated the relationship between remote cholecystectomy, development of AMR and PABSI outcomes. Several studies, however, have investigated the impact of cholecystectomy on *C. difficile* incident infection (CDI), recurrence and mortality [50, 51]. Given that cholecystectomy is associated with increased levels of fecal secondary bile acids which confer resistance to *C. difficile* infection in animal models, the authors proposed that cholecystectomy might protect against CDI. Patients without a gallbladder did not experience higher rates of incident CDI or recurrence. However, there was a significant increase in mortality in patients following cholecystectomy [51]. The reason for the increase in mortality is poorly understood. Conversely, in our study investigating PABSI, having a prior remote cholecystectomy did not impact clinical outcome as patients with and without a gallbladder had similar durations of bacteremia, in-hospital and 90-day mortality rates. There were baseline differences in the two study populations that may have masked outcome differences. Neutropenia, a known risk factor for poor outcomes in PABSI, and leukemia/lymphoma were more common in patients with a gallbladder, whereas peripheral vascular disease and liver disease (other characteristics associated with poor PABSI outcomes) were more common in patients without one. Furthermore, patients without a gallbladder experienced a longer delay in the onset of active antimicrobial therapy and averaged higher CCI scores at time of PABSI diagnosis. These differences may have offset and contributed to similar outcomes seen in both patient populations. Overall, given these confounders, it is difficult to determine the impact of baseline population differences on outcomes in our study.

Additionally, we found no association between prior cholecystectomy and increased antibiotic resistance among PABSI isolates as there were no statistically significant differences in AMR patterns dependent on the presence or absence of a gallbladder at the time of diagnosis. We had theorized that in patients with an intact gallbladder during PABSI, the *Pa* isolate may have increased bile exposure, a risk factor for the development of AMR [41]. Other pathogens such as *E. coli*, *Klebsiella pneumoniae*, and enterococci have displayed unexpected resistance traits when cultured from the gallbladder, and bile exposure has been shown to increase expression of efflux pumps critical for the development of antibacterial resistance [20, 34, 52, 53]. Thus, the lack of increased AMR in patients with a retained gallbladder was surprising. However, several factors may explain this observation. First, the quantity of bile exposure in patients following cholecystectomy is difficult to ascertain. Even in the absence of a gallbladder the liver produces bile which is subsequently excreted into the small intestine. Furthermore, patients who have undergone prior cholecystectomy have had at least one contact with the healthcare system, thereby increasing their exposure to healthcare-associated pathogens which routinely display higher patterns of antimicrobial resistance [54–57]. Finally, it is likely that in a percentage of our cohort, PABSI arose from sites other than the GI tract (e.g. from the lungs or exogenous via indwelling catheters). In these cases, it is unlikely that *Pa* would have been exposed to bile in either the gallbladder or GI tract prior to infection. These factors may have obscured any differences in AMR related to the presence or absence of a gallbladder.

This study was able to place PABSIs at our institution in historical context as a similar study, completed more than a decade earlier, assessed morbidity associated with PABSI. In that study, the average patient was 57 years old, the in-hospital mortality rate was 37% [9] and one-third of all infections were polymicrobial. By comparison, our cohort was older (average age 66.9 years), the in-hospital mortality was lower (20.8%), and only 17.3% of all bloodstream infections were polymicrobial. We are encouraged by the 16% decline in the in-hospital mortality rate of PABSI over the last 15 years and believe that there are several factors that have contributed to this decline. First, we believe that recognition of rising rates of AMR, particularly in *Pa* infections, has led to more rapid and aggressive initiation of anti-pseudomonal antibiotics early in the hospital stay in those patients with risk factors for *Pa* infections. Additionally, during the study period, newer combination antimicrobials for *Pa* infections including ceftazidime-avibactam and ceftolozane-tazobactam were introduced (2014 and 2015, respectively). Both have expanded the antibiotic toolkit available to treat MDR and XDR *Pa* infections. Lastly, there are differences in study populations between the two studies. Though our cohort was considerably older than the original study, there was a dramatic reduction in the rate of polymicrobial bloodstream infections. These factors may offset one another; however, our study is not powered to fully assess the impact of these differences on outcomes. Despite the 15-year interval between the two studies, it is important to acknowledge that the overall mortality rate of PABSI remains high.

General estimates suggest that the lifetime prevalence of cholecystectomy is approximately 4.0% overall, 5.3% among women and 2.4% among men [58]. In our cohort, 22.0% of our patients with PABSI had prior cholecystectomy. Contrary to our hypothesis that cholecystectomy may confer protection against PABSI, the disproportionate number of patients without a gallbladder suggests that this population may, in fact, be vulnerable to PABSI or have significant risk factors that predispose them to PABSI.

This study has several important limitations. First, the population size was relatively small and included patients at a single center, and generalizability to larger populations and different clinical settings may be limited. Additionally, the data collected reflected a disproportionate ratio (three to one) of patients with and without a gallbladder with overall rates of cholecystectomy that were five times higher than expected based on general estimates. A more balanced population and a matched cohort approach may serve to better highlight subtle differences. Secondly, this analysis was limited to the first episode of PABSI observed in each patient. As prior infection predisposes patients to *Pa* colonization and increases the risk of invasive infection, there may have been colonized individuals in the cohort who were more predisposed to invasive infection [15, 59]. This study did not assess whether cholecystectomy impacts *Pa* carriage in the gastrointestinal tract, an important risk factor for invasive disease. Bachta, et al. demonstrated that the gallbladder is crucial for fecal excretion of *Pa* in mice [19]; However, its role in amplification of fecal shedding in humans is unknown. Additionally, most patients received prompt active antimicrobial therapy within one day of their first positive blood culture (mean days to active therapy 0.73 ± 2.1 days), limiting our ability to assess associations between onset of appropriate antimicrobial therapy, underlying resistance patterns and outcomes.

While we did not observe an improvement in PABSI outcomes in patients lacking a gallbladder, we were able to observe improved overall survival rates following PABSIs compared to a prior study at our institution. This suggests that recognition and treatment of PABSI may be improving, but there is still much to learn about carriage, colonization and excretion of *Pa*. Given the observation that the gallbladder promoted increased gastrointestinal excretion of *Pa* in mice, it would be interesting to investigate this question in human patients. A future prospective study comparing fecal carriage of *Pa* in patients with and without a gallbladder following PABSI would help elucidate whether the gallbladder is important for amplification of *Pa* in patients. If the presence of a gallbladder provided an important *in vivo* replication niche and increased excretion of *Pa* this may change the way clinicians approach patients with disseminated *Pa* infections. Such patients might benefit from increased infection control measures that limit environmental contamination of their rooms. Additionally, prophylactic elective cholecystectomy in patients with recurrent BSI or a lower threshold for emergent cholecystectomy in critically ill patients could be entertained. Ultimately, it is critical that we continue to explore the interplay between the gallbladder, bile exposure and antimicrobial resistance in the context of PABSI to better understand the pathophysiology of BSIs and to enhance our ability to develop new therapeutics in light of rising rates of antimicrobial resistance.

## STATEMENTS AND DECLARATIONS

### Ethics approval and consent to participate

This research study (STU00210641) was reviewed by the Northwestern University Institutional Review Board and was deemed an observational study where no ethical approval was required.

### Authors’ contributions

The authors confirm contribution to the paper as follows: study conception and design: L.R.H., M.P.A., A.R.H., and K.E.R.B; data collection: H.K.B., L.R.H., and K.E.R.B.; analysis and interpretation of results: H.K.B., M.S., L.R.H., and K.E.R.B; draft manuscript preparation: H.K.B., M.S., and K.E.R.B. All authors reviewed the results and approved the final version of the manuscript.

## Data Availability

All data produced in the present study are contained within the manuscript.

## ABBREVIATIONS

(*Pa*): *Pseudomonas aeruginosa*
(PABSI): *Pseudomonas aeruginosa* bloodstream infection
(BSI): bloodstream infection
(AMR): antimicrobial resistance
(MDR): multidrug-resistant
(XDR): extensively drug-resistant
(GB): gallbladder
(GI): gastrointestinal

